# Probabilistic Forecasting of Monthly Dengue Cases Using Epidemiological and Climate Signals: A BiLSTM–Naive Bayes Model Versus Mechanistic and Count-Model Baselines

**DOI:** 10.1101/2025.10.20.25338419

**Authors:** Michael Marko Sesay, Antony Ngunyi, Herbert Imboga

**Affiliations:** Department of Mathematics, Pan African University, Institute for Basic Sciences, Technology and Innovation, Kiambu, Kenya; Department of Statistics and Actuarial Sciences, Dedan Kimathi University of Technology, Nyeri, Kenya; Department of Statistics and Actuarial Sciences, Jomo Kenyatta University of Agriculture and Technology, Kiambu, Kenya

## Abstract

Reliable short-term forecasts can help urban health systems anticipate dengue surges and allocate resources. We assembled monthly dengue case counts for Freetown, Sierra Leone (2015–2025), and compared four probabilistic model families under a leakage-safe, rolling-origin protocol at 1–3-month horizons: a negative-binomial generalized linear model (NB-GLM), a negative-binomial INGARCH, a mechanistic renewal model with NB observations, and a Bidirectional LSTM with a negative-binomial output (BiLSTM-NB). All models used the same seasonal harmonics and autoregressive lags; “light” climate inputs (rainfall, temperature, relative humidity) were limited to ≤ 3 features and lagged to reflect real-time availability. Evaluation used proper mean log score, coverage, median width of 50% and 90% predictive intervals, randomized PIT histograms, and Diebold–Mariano tests with Newey–West standard errors. For the main comparison, we aligned forecasts on common (issue, target) pairs per horizon (n=33). No single approach dominated across horizons. At 1–2 months, the INGARCH-NB and NB-GLM frequently achieved the highest mean log scores with near-nominal coverage. At 3 months, a calibrated BiLSTM-NB ensemble more often yielded the best mean log score and narrower central intervals without marked undercoverage. Diebold–Mariano tests indicated statistically significant improvements for INGARCH-NB over mechanistic renewal at shorter horizons and for BiLSTM-NB over mechanistic and GLM baselines at 3 months. PIT profiles were closer to uniform for the top-performing model at each horizon, supporting probabilistic calibration.These results suggest a horizon-specific toolkit for operational dengue forecasting: lean count models for 1–2 months ahead and calibrated deep learners for 3 months, all under consistent leakage controls. The framework and artifacts (per-issue forecasts, aligned indices, and code) provide a transparent baseline for future work in similar urban settings.

**Author summary:** We conducted this study to help public health teams in Freetown, Sierra Leone, plan clinical capacity and vector control using short-term dengue forecasts they can trust. Many forecasting approaches exist, but they are rarely compared under the same, leakage-safe conditions. We assembled monthly dengue case data (2015–2025) and created consistent seasonal and autoregressive features for all models, using only a light, real-time-feasible set of climate inputs. We then compared four model families: a negative-binomial generalized linear model, an INGARCH count model, a mechanistic renewal model, and a Bidirectional LSTM with a negative-binomial output. Using an expanding-window, rolling-origin evaluation at 1-3 month horizons, we assessed proper scoring, interval coverage and width, PIT histograms, and Diebold–Mariano tests on aligned targets. We found that no single method dominated across horizons: simpler count models performed strongly at 1-2 months, while a calibrated BiLSTM provided the best performance more often at 3 months. These findings suggest a horizon-specific toolkit can improve operational dengue forecasting in similar urban settings.

## Introduction

Dengue fever remains one of the most pervasive vector-borne diseases worldwide, affecting tropical and subtropical regions across the globe with an expanding footprint. The geographic spread of dengue continues to increase, driven by recurring outbreaks that contribute to significant morbidity and place a growing burden on public health systems. This expansion is partly attributable to complex interactions among various factors, including climatic changes that alter mosquito breeding habitats and virus survival and rapid urbanization that increases human-mosquito contact zones. These factors collectively exacerbate transmission dynamics, triggering more frequent outbreaks and posing new challenges for disease control. Given these complexities, there is a compelling operational need to develop reliable, short-term forecasting tools that assist health authorities in anticipating dengue incidence, thus enhancing preparedness and response capacities globally [1–3].

Among regions grappling with emerging dengue risks, West Africa presents unique epidemiological characteristics that call for focused attention. Increasingly, evidence has highlighted sustained local transmission of dengue, dispelling earlier assumptions of sporadic or imported cases. The region features complex ecological and socio-economic factors, such as seasonal rainfall patterns, temperature variability, and urban growth, that influence mosquito population dynamics and virus transmission potential. This evolving landscape underscores the necessity for improved situational awareness through enhanced surveillance and data-driven forecasting. Regional efforts to understand dengue epidemiology are growing in importance, as timely, region-specific information is crucial for mobilizing interventions and curbing outbreaks that impose considerable health and economic burdens on vulnerable populations [4–6].

Focusing more narrowly, Freetown, the capital of Sierra Leone, represents a pertinent case study for operational dengue forecasting owing to its routine monthly surveillance data and documented dengue activity. This city’s epidemiological context, characterized by periodic outbreaks and climatic conditions favorable for vector proliferation, creates a pressing need for targeted forecasting approaches to inform public health decision-making. Monthly forecasting, in particular, serves as a pragmatic temporal resolution balancing data availability and operational utility. It supports health officials in anticipating case surges, allocating resources efficiently, and deploying vector control measures in a timely fashion. The availability of consistent data streams allows the construction of models attuned to local transmission dynamics, fostering actionable insights for outbreak preparedness and ultimately contributing to the reduction of dengue’s impact in Sierra Leone [7, 8].

Monthly dengue counts exhibit overdispersion, strong annual seasonality with short serial dependence, and complex relationships with environmental drivers, challenging traditional time series approaches [9, 10]. Negative binomial generalized linear models (NB-GLMs) offer interpretable covariate effects and handle overdispersion but may inadequately capture temporal feedback dynamics [11, 12]. NB-INGARCH models explicitly incorporate dependence of the conditional mean on both past observations and past conditional means, providing observation-driven volatility modeling for count data [13, 14]. Renewal models relate incidence to a time-varying reproduction number (*R*_*t*_) and serial-interval kernel, supporting epidemiological interpretation while maintaining parsimony [15]. Modern BiLSTM architectures with negative-binomial output heads learn complex non-linear patterns while yielding calibrated probabilistic count forecasts [16]. However, time-series machine learning remains susceptible to information leakage through improper feature construction and validation design, requiring careful feature timing and rolling-origin evaluation protocols.

Despite extensive methodological development, leakage-safe, aligned comparisons of count baselines, observation-driven, mechanistic, and deep sequence models for monthly dengue forecasting in West Africa remain limited [17]. We address this gap with an aligned, expanding-window evaluation in Freetown comparing NB-GLM, INGARCH-NB, Renewal-NB, and a BiLSTM-NB architecture incorporating autoregressive skip connections and isotonic calibration. We analyze monthly reported dengue cases in Freetown (2015–2025) with monthly rainfall, air temperature, and relative humidity aggregates as potential environmental drivers. All models include month harmonic features and autoregressive lags ({*y*_*t*−1_, *y*_*t*−2_, *y*_*t*−3_, *y*_*t*−12}_) to capture both seasonal and short-term dependencies. Baseline models employ a light set of ≤ 3 climate features when real-time availability permits, while the BiLSTM uses climate variables exclusively at lag-1 to prevent data leakage.

We employ an expanding-window rolling-origin protocol at horizons *h* ∈ { 1, 2, 3 } months ahead, with a minimum training length of 48 months to ensure adequate seasonal pattern estimation. Evaluation emphasizes mean log score as a strictly proper scoring rule, central 50% and 90% prediction interval coverage, median widths for calibration and sharpness assessment, randomized PIT histograms for distributional calibration, and Diebold-Mariano tests with Newey-West standard errors on aligned issue-target indices [18]. Our contributions include:

- Leakage-safe feature timing (lag-1 climate restriction), seed-ensemble BiLSTM-NB with autoregressive skip connections, and optional isotonic calibration for improved reliability
- Head-to-head comparison of NB-GLM (direct forecasting), INGARCH-NB (observation-driven), Renewal-NB (mechanistic), and BiLSTM-NB under shared seasonal and autoregressive structure
- Aligned backtesting enabling fair Diebold-Mariano model pairs, with unaligned results preserved in supplementary materials
- Operational metrics emphasizing proper scoring rules, reliability, and sharpness for public health decision support

The remainder of this paper is organized as follows: Section 2 reviews related work in dengue forecasting and count time series modeling. Section 3 describes data sources, feature engineering, model formulations, experimental setup, and evaluation metrics. Section 4 presents comparative results, diagnostic analyses, and robustness checks. Section 5 discusses implications for operational dengue forecasting in resource-limited settings and identifies future research directions.

## Materials and methods

### Data Descriptions

This study utilizes a curated dengue surveillance and climate dataset covering the period from January 2015 to December 2024 for Freetown, Sierra Leone. The dataset integrates epidemiological records of reported dengue cases with key climatic variables to support time series forecasting using deep learning techniques. The dengue case data represent the monthly number of laboratory-confirmed and clinically suspected dengue infections in Freetown. The data were collated and preprocessed to ensure temporal continuity, with missing months imputed where appropriate. Each observation corresponds to the total number of dengue cases recorded in a given month.

The climatic variables were obtained from publicly available meteorological sources. The selected features include. Temperature (^°^*C*): Monthly average surface temperature. Humidity (%): Monthly average relative humidity. Precipitation (mm): Monthly total accumulated rainfall. These climatic variables were selected because they are known to influence the population dynamics of the *Aedes* mosquito vector and the transmission intensity of dengue. An exploratory analysis was conducted to assess temporal trends and seasonality. Figure 1b presents the average monthly dengue case profile over the study period. The results indicate that transmission peaks typically occur between June and September, coinciding with the rainy season, while the lowest transmission occurs during the dry months (December–April).

**Fig 1.**
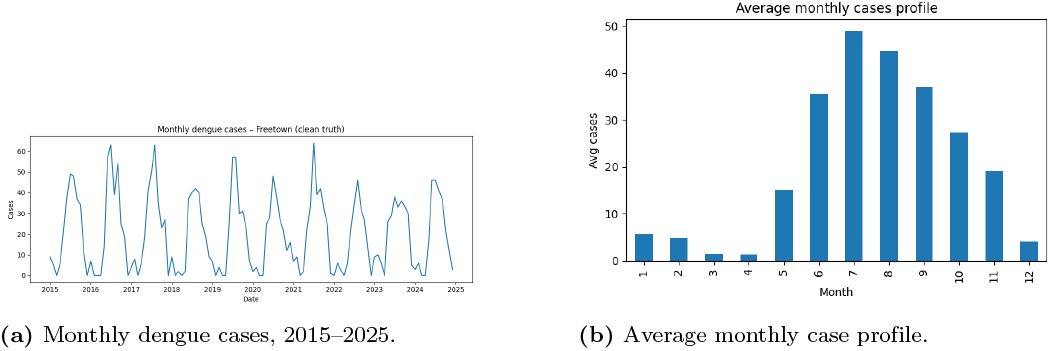
Monthly dengue in Freetown. (A) Monthly reported cases from 2015 to 2025, showing clear seasonality and recurrent annual peaks. (B) Average monthly case profile highlighting the seasonal transmission pattern.

### Data Preprocessing and Feature Engineering

Daily/weekly reports are timestamped, mapped to the first day of their calendar month, and aggregated to monthly counts. Let *Y*_*t*_ ∈ ℕ_0_ denote the number of reported dengue cases in month *t*. Non-numeric entries are coerced or dropped, and negative values are not permitted. Months with missing totals are retained as missing responses and excluded only from likelihood contributions that require them; feature construction (e.g., lags) respects missingness by truncating the applicable window. To capture annual structure, we construct 12–month trigonometric harmonics from the calendar month *m*_*t*_ ∈ { 1, …, 12 }:

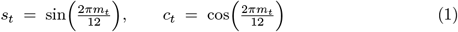

These harmonics enter all models as external regressors and are standardized within each training fold (below). Short- and medium-range dependence is represented by integer lags of the case series,

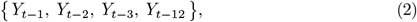

which are available to all models. When a required lag is unavailable (e.g., at the beginning of the series), the corresponding issue month is omitted from training/forecasting for models that need that lag. Environmental drivers are limited to a “light” set of at most three aggregate climate variables: monthly rainfall, near-surface air temperature, and relative humidity. To prevent look-ahead:

- For mechanistic and count baselines (NB–GLM, INGARCH–NB, Renewal–NB), we allow either contemporaneous (*t*) or lag–1 (*t* − 1) values in accordance with *real–time* availability in an operational setting; rainfall is used at lag–1 by design.
- For the BiLSTM–NB, *all* climate inputs are strictly lagged by one month, i.e., 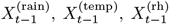, to provide a conservative guard against leakage.

Let *x*_*t*_ denote the vector of selected climate covariates at month *t*. Climate covariates are standardized within each training fold by

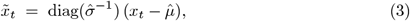

where 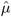 and 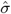 are the mean and standard deviation computed *only* on the current training window; these statistics are then applied to the corresponding validation/test issues. Trigonometric harmonics (*s*_*t*_, *c*_*t*_) are scaled analogously. Count lags { *Y*_*t*−𝓁_ } are left unscaled. For recurrent models, we form leakage-safe supervised sequences with a fixed lookback *W* = 12 months. The count stream for issue *t* is

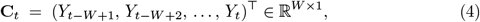

and the auxiliary feature vector is

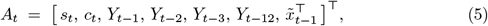

with climate at lag–1 only. Only issues for which all required elements of **C**_*t*_ and *A*_*t*_ are present are used for training and evaluation. We construct multi–step targets for *h* = 1, 2, 3 months ahead,

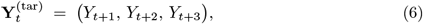

which are paired with inputs (**C**_*t*_, *A*_*t*_) for direct multi–horizon training (BiLSTM–NB) or for per–horizon generalized linear fits (NB–GLM). In all cases, targets *Y*_*t*+*h*_ are never used in feature computation at issue *t*. All feature engineering for issue *t* uses information available at or before *t* (and calendar month indices for *t*+*h* where needed). Standardization parameters are recomputed for each expanding training fold and *not* informed by validation/test data. This protocol ensures that model fitting and scoring mimic the information constraints of real-time forecasting. We persist the per–issue (`issue_date, target_date`) keys and the training–fold scaling statistics used at each issue. These artifacts allow exact regeneration of the aligned evaluation sets and facilitate audit of leakage safeguards.

### Models

All models produce probabilistic forecasts for monthly counts using a Negative-Binomial (NB2) observation model with mean *µ* and dispersion *α*, where Var(*y*|*µ*) = *µ* + *α* · *µ*^2^. Except where noted, we fit separate direct models per horizon (*h*).

### NB-GLM

The negative binomial generalized linear model (NB-GLM) extends the Poisson GLM to accommodate overdispersion commonly observed in dengue counts [11, 12]. We adopt the NB2 mean-variance relationship. For horizon *h* ∈ {1, 2, 3}, the monthly count *Y*_*t*+*h*_ conditional on the information set ℱ_*t*_ is modeled as

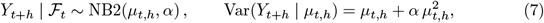

where *µ*_*t,h*_ = 𝔼 [*Y*_*t*+*h*_ | ℱ_*t*_] and *α >* 0 is the overdispersion parameter [19]. As *α* → 0 the model approaches Poisson; larger *α* implies greater overdispersion [20]. For likelihood computations we use the (*r, p*) parameterization

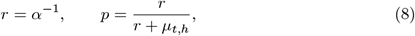

with pmf

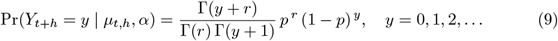

We use a *horizon-specific* linear predictor (direct strategy) [21]:

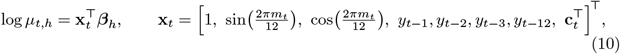

where *m*_*t*_ is the calendar month, and **c**_*t*_ is a *light* climate vector with at most three covariates chosen from rainfall, air temperature, and relative humidity. To prevent leakage, climate terms are restricted to lag-1 (e.g., rainfall_*t*−1_); if an operational pipeline provides same-month readings at issue time, contemporaneous terms are allowed only when available at *t* (never *t* + *h*). Seasonal harmonics and climate are standardized on the *training folds only* ; count lags remain unscaled. Let ℐ_*h*_ = { *t*_0_, …, *T* − *h* } be the set of training indices after respecting the maximal lag and minimum training length. With *r* = *α*^−1^ and 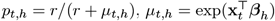, the log-likelihood is

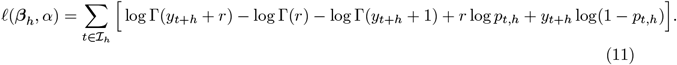

We estimate (***β***_*h*_, *α*) by alternating (i) IRLS updates for ***β***_*h*_ under an NB2 working model and (ii) a Pearson method-of-moments update for *α*:

#### 1. IRLS for *β* (given *α*^(*k*)^)

Let 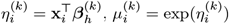. The NB2 working weight and response are

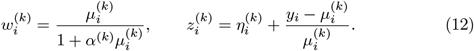

The weighted least-squares update is

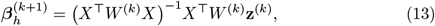

with *X* the design matrix, 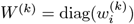, and 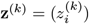.

#### 2. MoM for *α* (given 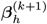)

With 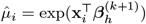 and Pearson residuals 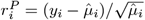, use 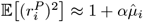 Regress 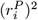 on 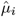 with intercept and take the slope as 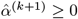.

Iterate until 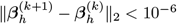 (and negligible change in *α*). All features in **x**_*t*_ are computed from information available at issue time *t*; standardization parameters are learned on training folds and applied to validation/test folds.

For a new issue *T*,

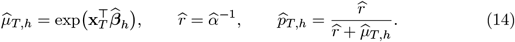

Point forecasts are 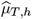. Central (1 − *τ*) × 100% prediction intervals use NB quantiles

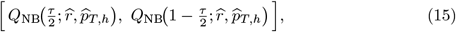

e.g., τ = 0.5 (50%) and τ = 0.1 (90%)

### INGARCH-NB

The integer-valued GARCH-type model with negative-binomial innovations (INGARCH-NB) adapts volatility-style feedback to count data, capturing short-memory dependence and overdispersion frequently observed in dengue surveillance series [13, 22]. We adopt the NB2 mean–variance form to maintain consistency with all baselines.

Let ℱ_*t*−1_ = *σ*(*Y*_*t*−1_, *Y*_*t*−2_, …) be the natural filtration. We assume

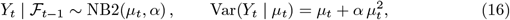

with overdispersion *α >* 0. For likelihood evaluation, we map to the (*r, p*) parameterization, *r* = *α*^−1^ and *p* = *r/*(*r* + *µ*_*t*_), with pmf 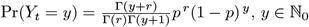.

We use a log link with seasonal harmonics, an observed-count feedback, and a conditional-mean feedback:

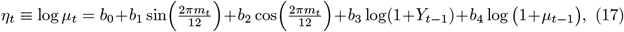

so that *µ*_*t*_ = exp(*η*_*t*_). The log(1 + ·) transform stabilizes the feedback at zero counts and avoids numerical issues [14]. The term in log(1 + *µ*_*t*−1_) provides GARCH-type persistence in the conditional mean. Seasonal harmonics { sin, cos } capture annual dengue cyclicality. (Climate regressors are *not* included in the INGARCH specification in our main analysis to keep the mechanistic lift attributable to dynamics rather than exogenous inputs.)

Given *y*_1:*T*_ and an initialization *µ*_0_ *>* 0, the log–likelihood is

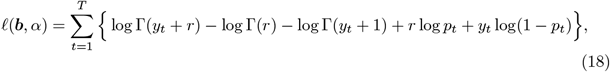

where *r* = *α*^−1^, *p*_*t*_ = *r/*(*r* + *µ*_*t*_), and *µ*_*t*_ is generated recursively from (17). The recursion requires sequential evaluation of *µ*_*t*_(***b***, *α*) for *t* = 1, …, *T*.

We compute the maximum likelihood estimator 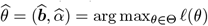 using box–constrained quasi–Newton (L–BFGS–B). To promote stability we constrain *α >* 0 and *b*_4_ ∈ [0, 1), the latter limiting explosive feedback in the *µ*_*t*−1_ channel (heuristic stationarity control in the log domain). Gradients can be propagated through the recursion by the chain rule:

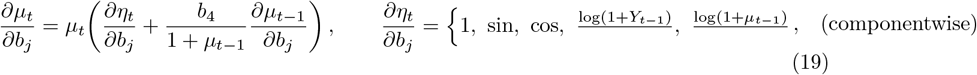

and ∂𝓁/∂*µ*_*t*_ obtained from (18). In practice, finite–difference gradients with a well–chosen initialization (e.g., 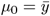) are adequate.

### Renewal–NB

We adopt an epidemiological renewal model with a negative binomial (NB2) observation process as a mechanistic baseline for monthly dengue forecasting. The formulation decomposes incidence into an effective reproduction component and a weighted sum of past infections, cohering with standard epidemic renewal theory [15, 23].

Let ℱ_*t*−1_ denote the information set up to month *t* − 1. Counts follow

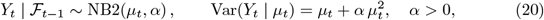

with negative-binomial parameters mapped as *r* = *α*^−1^ and *p*_*t*_ = *r/*(*r* + *µ*_*t*_); the pmf is 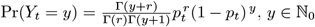.

The discrete renewal equation generates monthly incidence

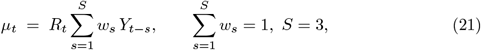

where *w* = (*w*_1_, *w*_2_, *w*_3_) ∝ (0.6, 0.3, 0.1) encodes a short generation interval at a monthly scale (mass concentrated at one to two months), consistent with dengue intrinsic and extrinsic incubation plus reporting delay [24]. The effective reproduction number is seasonally modulated,

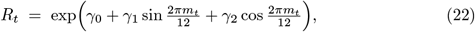

with *m*_*t*_ ∈ {1, …, 12} the calendar month. This ensures *R*_*t*_ *>* 0 and allows seasonal forcing around the baseline exp(*γ*_0_). (Interpretation: *R*_*t*_ *>* 1 growth, *R*_*t*_ = 1 equilibrium, *R*_*t*_ *<* 1 decline.)

A light climate variant augments (22) during *fitting* only,

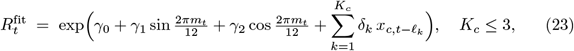

where 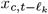 are climate covariates (rainfall, temperature, relative humidity) at honest lags (no *t* + *h* information). For *forecasting*, we revert to the seasonal form (22) to avoid reliance on future climate, thereby eliminating look-ahead risk. This “fit-with-climate, forecast-seasonal” strategy isolates mechanistic lift from seasonality while respecting real-time constraints.

Given *y*_1:*T*_ and *S* = 3, the renewal recursion is well-defined from *t* = *S* + 1. The log-likelihood under NB2 is

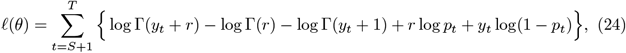

with *θ* = (*γ*_0_, *γ*_1_, *γ*_2_, *α*), *r* = *α*^−1^, *p*_*t*_ = *r/*(*r* + *µ*_*t*_), and *µ*_*t*_ given by (21)–(22) (or (23) during climate-augmented fitting).

We obtain the 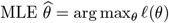 via box-constrained L–BFGS–B. Practical constraints: *α >* 0; a soft bound 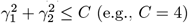 to prevent unrealistically large seasonal amplitudes. Analytical gradients follow from

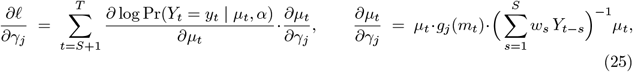

where *g*_0_ ≡ 1, *g*_1_ ≡ sin(2*πm*_*t*_*/*12), *g*_2_ ≡ cos(2*πm*_*t*_*/*12). In practice, finite-difference derivatives with careful scaling are sufficient due to the low parameter dimension.

Let *T* be the last observed month. For *h* = 1,

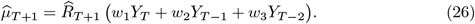

For *h >* 1.

### BiLSTM-NB

The Bidirectional Long Short–Term Memory model with a Negative–Binomial output head (BiLSTM–NB) couples deep sequence representations with a count–likelihood tailored to overdispersed dengue surveillance data. It learns non-linear dependencies while producing horizon-specific predictive distributions suitable for probabilistic evaluation [25, 26].

At monthly cadence, each training instance at issue time *t* comprises (i) a univariate count window

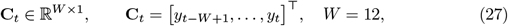

and (ii) an auxiliary vector

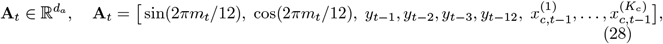

where *m*_*t*_ is the calendar month; the AR lags are fixed as (1, 2, 3, 12); and the “light” climate set uses up to *K*_*c*_ ≤ 3 lag-1 features among rainfall, temperature, and relative humidity. To prevent look–ahead bias, (a) only lagged climate is used, (b) harmonics and climate are standardized on training folds only, and (c) rolling-origin backtesting uses expanding windows (minimum 48 months). The count window is passed through two stacked bidirectional LSTM layers with 32 units each. Denote the final sequence embedding by **h**_BiLSTM_ ∈ ℝ^64^. A dense block with ReLU and dropout (rate 0.2) produces a non-linear summary **h** ∈ ℝ^64^. To retain short–memory linear structure, we include an *autoregressive skip* that maps the four AR lags to horizon-specific logits. The network then concatenates the dense representation with auxiliary features,

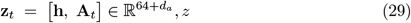

and outputs, for each horizon *h* ∈ { 1, 2, 3 }, two real numbers that are transformed into the NB2 parameters.

For horizon *h*, the model outputs pre-activations 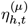 and 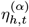 via affine maps of **z**_*t*_ plus an AR-skip term for the mean,

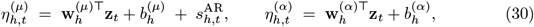

where 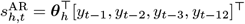. Positivity and numerical stability are enforced with bounded activations:

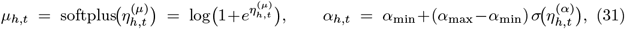

with *α*_min_ = 10^−4^, *α*_max_ = 2, and *σ*(·) the logistic sigmoid [27]. We adopt NB2 with mean *µ*_*h,t*_ and variance 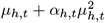 in (*r, p*) form, *r*_*h,t*_ = 1*/α*_*h,t*_ and *p*_*h,t*_ = *r*_*h,t*_*/*(*r*_*h,t*_ + *µ*_*h,t*_).

Let *y*_*t*+*h*_ be the target for horizon *h*. The per-instance, multi-horizon negative log-log-likelihood is

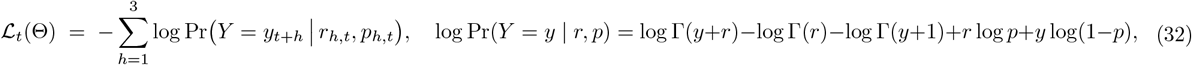

and the empirical loss averages ℒ_*t*_ over training indices. We optimize with Adam (learning rate 10^−3^), gradient–norm clipping (∥∇∥ ≤ 1.0), early stopping (patience = 12 epochs), and `ReduceLROnPlateau` (factor 0.5, patience = 6).

Bidirectional LSTMs extract seasonally phased patterns and non-linear lag effects; the AR skip preserves sharp short-term responses often attenuated by deep encoders; the NB2 head matches the observed overdispersion; bounded *α* prevents pathological variance; ensembling stabilizes small-sample training; and isotonic calibration addresses minor miscalibration frequently observed in neural density estimators [27]. Altogether, this yields a leakage–safe, probabilistic forecaster aligned with operational constraints.

### Experimental Setup

We evaluate all models under an expanding-window, rolling-origin design for monthly dengue surveillance (study window: 2015–2025). Let *t* index months and 𝒟_1:*t*_ denote data up to and including *t*. After a minimum of 48 training months and once all required features exist, each model is (re)fit on 𝒟_1:*t*_ and issues *h*-step–ahead probabilistic forecasts for *h* ∈ { 1, 2, 3 }, targeting (*t*+*h*). This procedure repeats for every eligible issue month, yielding per–issue predictive distributions and realized outcomes for scoring. For every model and eligible (*t, h*), we record the predictive NB parameters 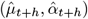, full predictive distribution (via 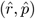 with 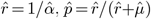, the realized *Y*_*t*+*h*_, log score log *P* (*Y*_*t*+*h*_ ℱ_*t*_), central 50% and 90% intervals and their coverages, and randomized PIT values. These per-issue records underpin the summary metrics and Diebold-Mariano tests in Section.

### Evaluation Metrics

We employ a comprehensive suite of metrics to assess both probabilistic accuracy and calibration of forecasts, following best practices for count data evaluation [28, 29].

#### Mean Log Score

The logarithmic scoring rule serves as our primary metric for probabilistic forecast quality, providing a strictly proper scoring rule that incentivizes honest reporting of predictive distributions [29]. For a forecast with negative binomial distribution parameters 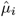 and 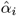, and observed value *y*_*i*_, the log score is:

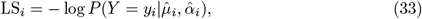

where the negative binomial probability mass function is:

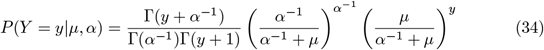

The mean log score across *n* forecasts is:

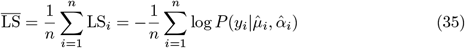

Lower scores indicate superior probabilistic accuracy. The log score heavily penalizes forecasts that assign low probability to observed outcomes, making it particularly sensitive to outlier predictions [30].

#### Predictive Interval Coverage

Calibration assessment employs coverage rates of central prediction intervals at nominal levels *τ* ∈ { 0.50, 0.90 }. For each forecast *i*, we construct the equal-tailed prediction interval:

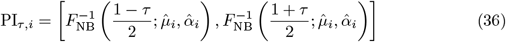

where 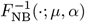 denotes the negative binomial quantile function. The empirical coverage rate is:

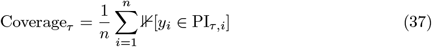

where ⊮[·] is the indicator function. Well-calibrated forecasts yield Coverage_*τ*_ ≈ τ . Systematic deviations indicate miscalibration: undercoverage (Coverage_*τ*_ *< τ*) suggests overconfidence, while overcoverage indicates excessive uncertainty [31].

#### Median Interval Width

Forecast sharpness, conditional on calibration, is quantified via the median width of prediction intervals:

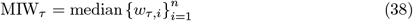

where the interval width for forecast *i* is:

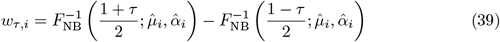

The median provides a robust measure less influenced by outliers than the mean. Among calibrated forecasts (those achieving nominal coverage), smaller MIW indicates sharper, more informative predictions [32]. We report MIW for both 50% and 90% prediction intervals to assess sharpness across different uncertainty levels.

#### Diebold-Mariano Test

Pairwise forecast comparisons employ the Diebold-Mariano (DM) test for equal predictive accuracy [18]. For two competing forecasts with log scores LS_1,*t*_ and LS_2,*t*_, define the loss differential:

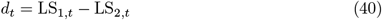

Newey-West

The null hypothesis *H*_0_ : 𝔼 [*d*_*t*_] = 0 (equal predictive accuracy) is tested using the statistic:

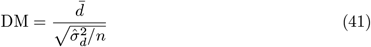

where 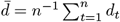 and 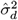 is the Newey–West heteroskedasticity and autocorrelation consistent (HAC) variance estimator is

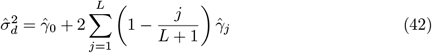

with autocovariances 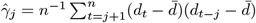 and bandwidth *L* = *h* – 1 to account for forecast error autocorrelation at horizon *h* [33]. Under *H*_0_ and standard regularity conditions, 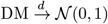. We employ two-sided tests at significance level *α* = 0.05, with negative values favoring model 1 and positive values favoring model 2 [34].

#### Probability Integral Transform (PIT) Histograms

Global calibration assessment utilizes probability integral transform (PIT) analysis. For continuous distributions, the PIT values 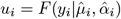 follow Uniform(0, 1) under perfect calibration. However, for discrete distributions like the negative binomial, standard PIT values exhibit discreteness artifacts [28].

We employ randomized PIT values to restore uniformity under calibration:

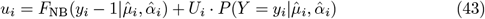

where *U*_*i*_ ∼ Uniform(0, 1) are independent random variables, and *F*_NB_(*y* − 1|*µ, α*) = *P* (*Y < y*|*µ, α*) with the convention *F*_NB_(−1|*µ, α*) = 0. This randomization ensures that *u*_*i*_ ∼ Uniform(0, 1) when forecasts are calibrated [35]

PIT histograms visualize the empirical distribution of 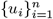 using 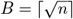 equal-width bins. Deviations from uniformity indicate specific calibration failures:

- U-shaped: Underdispersed forecasts (variance too small)
- Inverse-U shaped: Overdispersed forecasts (variance too large)
- Left-skewed: Systematic overforecasting (forecasts too high)
- Right-skewed: Systematic Anderson-Darling (forecasts too low)

We supplement visual inspection with the Anderson-Darling test for uniformity, though formal tests may have low power for small samples [36]. Consistency bands at level 1 − *α* are computed as

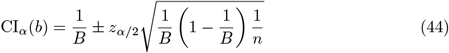

where *z*_*α/*2_ is the standard normal quantile and *b* ∈ {1, …, *B*} indexes bins [37].

### Results

We evaluated monthly probabilistic forecasts of dengue cases in Freetown, Sierra Leone, from 2015–2025 across three horizons *h* ∈ { 1, 2, 3 }. The candidate models comprise a count-regression baseline (NB-GLM), a light climate-augmented variant

(NB-GLM+Climate), a dynamic count model (INGARCH-NB), a mechanistic renewal model (Renewal-NB) with a seasonal *R*_*t*_ and a light climate-informed fit, and a deep sequence model (BiLSTM-NB). Performance was assessed using: (i) mean log score (primary), (ii) empirical coverage of nominal 50% and 90% predictive intervals (PIs), and (iii) median PI width (sharpness). To ensure fair comparison, all headline numbers are reported on *aligned* issue/target indices shared across models within each horizon (common *n* = 33); unaligned summaries are provided in the supplementary. Figures 2 visualize mean log scores and PI coverages, and Table 1 compiles the full summary.

**Table 1.**
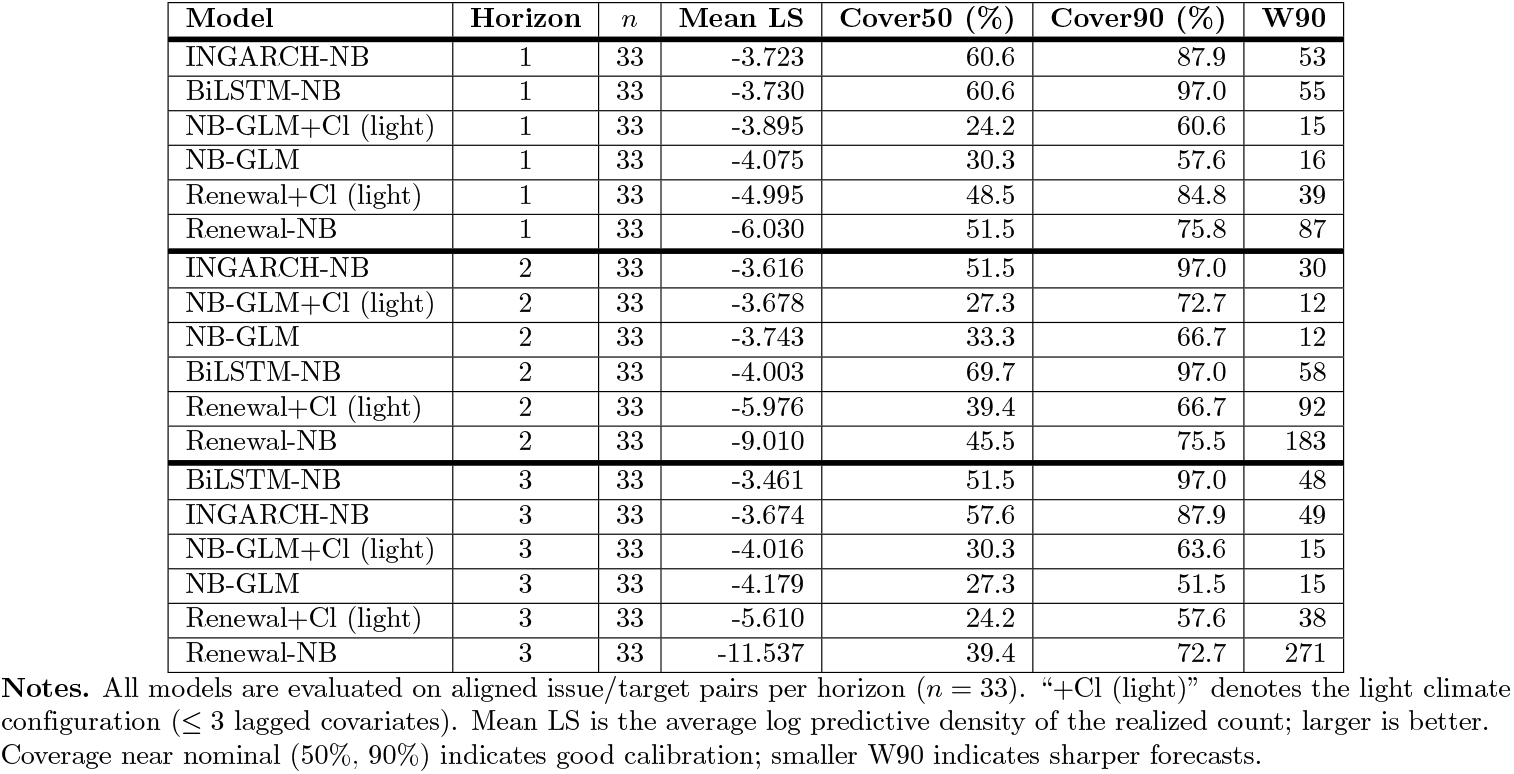
Aligned probabilistic performance by horizon (Freetown, monthly; 2015–2025). Higher mean log score (LS) is better. Coverage is empirical percentage; W90 is the median width of 90% prediction intervals (cases).

**Fig 2.**
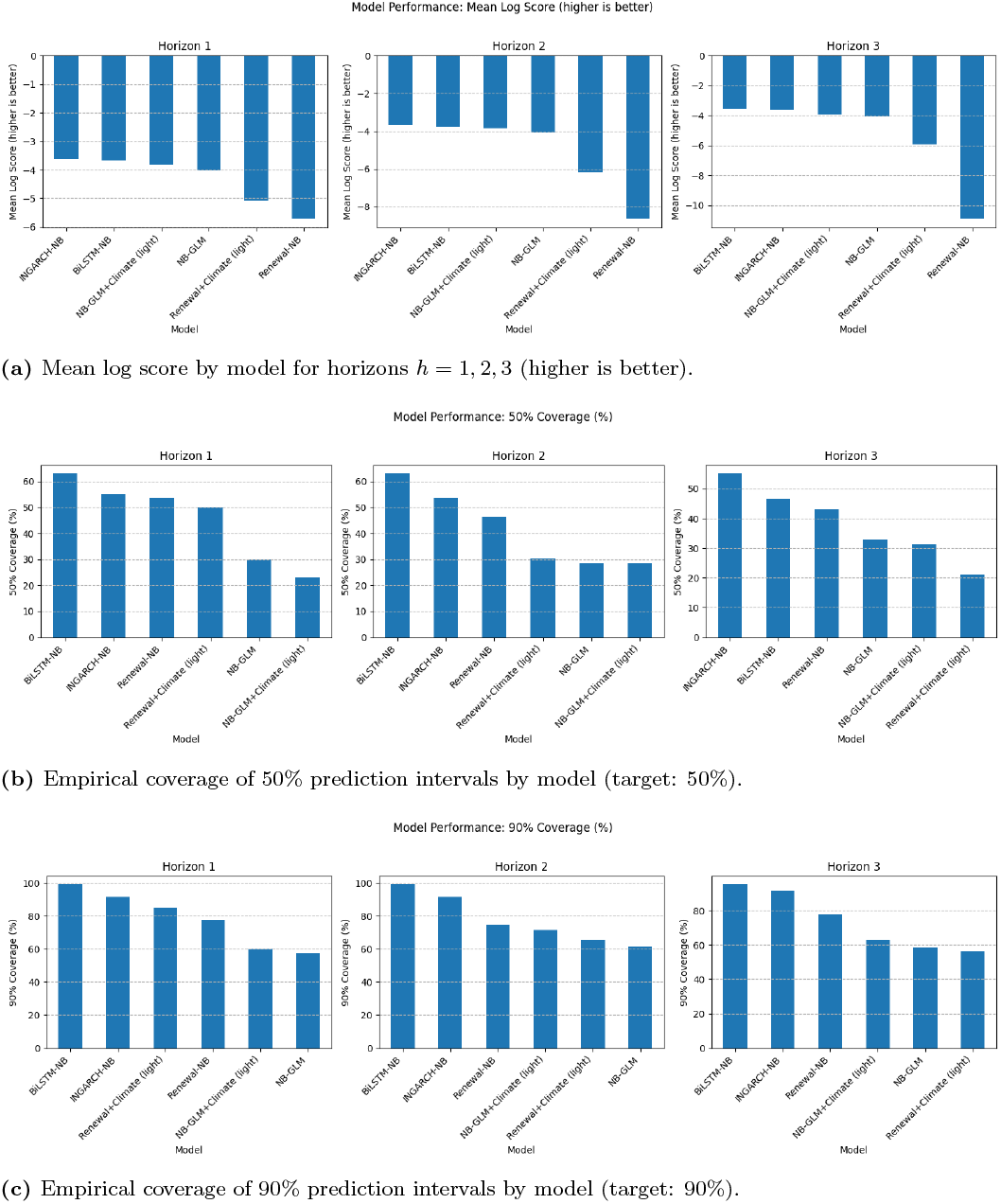
Probabilistic performance by model and horizon. Each row summarizes one metric across models: (A) mean log score (primary accuracy metric; higher is better); empirical coverage of 50% central prediction intervals; and (C) empirical coverage of 90% central prediction intervals (targets shown in captions).

#### Overall performance summary

At 1-month ahead, INGARCH-NB achieved the best mean log score (−3.723), followed closely by BiLSTM-NB (−3.730); both outperformed NB-GLM variants and Renewal models. At 2-months ahead, INGARCH-NB remained best (−3.616), followed by NB-GLM+Climate (−3.678) and NB-GLM (−3.743); BiLSTM-NB trailed here (−4.002). At 3-months ahead, BiLSTM-NB was best (−3.461), improving on INGARCH-NB (−3.674) and clearly ahead of NB-GLM variants (−4.016, −4.179) and Renewal models (−6.556, −10.648).

BiLSTM-NB produced near-nominal 90% coverage across horizons (97.0% at *h* = 1, 2, 3) with 50% coverage close to nominal (60.6%, 69.7%, 51.5%). INGARCH-NB showed reasonable calibration (50%: 60.6%, 51.5%, 57.6%; 90%: 87.9%, 97.0%, 87.9%). NB-GLM and NB-GLM+Climate consistently under-covered (e.g., 90% coverage at *h* = 3: 51.5% and 63.6%), indicative of overconfident intervals. Renewal models exhibited mixed calibration with broad intervals (see sharpness below).

NB-GLM variants were the sharpest (e.g., W90 = 15 at *h* = 1 and *h* = 3) but at the cost of under-coverage. BiLSTM-NB struck a balanced trade-off (W90 = 54, 58, 48 at *h* = 1, 2, 3 with high 90% coverage). Renewal models were least sharp, especially at *h* = 2 and *h* = 3 (e.g., Renewal-NB W90 = 201, 265).

##### Short vs. long horizons

At *h* = 1, INGARCH-NB and BiLSTM-NB are essentially tied on mean log score, both well-calibrated at 50% and with high 90% coverage. At *h* = 2, INGARCH-NB leads; BiLSTM-NB offers higher 50% coverage (69.7%) and near-nominal 90% coverage (97.0%) but at wider W90 (58). At *h* = 3, BiLSTM-NB is clearly best on mean log score while preserving high 90% coverage (97.0%) and competitive sharpness (W90 = 48).

##### Effect of climate features

Light climate improves NB-GLM at every horizon (e.g., mean log score improves from −4.075 to −3.895 at *h* = 1 and from −4.179 to −4.016 at *h* = 3), but these models still under-cover at 90%.

##### Operational implications

For near-term (*h* = 1) decision-making, either INGARCH-NB or BiLSTM-NB yields strong probabilistic accuracy; at medium range (*h* = 2) INGARCH-NB is preferable on accuracy; and for planning at longer range (*h* = 3) BiLSTM-NB provides the most informative probabilistic forecasts while maintaining excellent calibration.

Figure 2a displays mean log scores by model and horizon, while Figures 2b and2c show empirical coverage for nominal 50% and 90% PIs, respectively. The plots highlight three robust patterns: (i) INGARCH-NB dominates at *h* ∈ { 1, 2 } and BiLSTM-NB leads at *h* = 3; (ii) NB-GLM variants are the sharpest but under-cover; and (iii) Renewal-NB yields broad intervals with lower probabilistic accuracy. These visual trends are consistent with the summary statistics in Table 1.

#### Statistical significance of differences

To assess whether observed differences in probabilistic accuracy are more than sampling noise, we applied the two–sided Diebold–Mariano (DM) test on the *aligned* evaluation sets (common issue/target pairs per horizon; *n* = 33)Table 2. The loss differential was defined as the difference in log predictive density (log score) between two models for the same target; Newey–West standard errors used lag *L* = *h* − 1 to accommodate *h*-step dependence. Small *p*-values (*<* 0.05) indicate statistically significant differences in mean log score.

**Table 2.** Selected Diebold–Mariano tests on aligned samples (log score). Positive *t* favors Model 1; *p <* 0.05 indicates a statistically significant difference. We report only head-to-heads versus BiLSTM–NB and one anchor baseline comparison.

| Model 1 | Model 2 | Horizon | Common $n$ | $t$ | $p$ | Better |
| --- | --- | --- | --- | --- | --- | --- |
| **H=1 month** |  |  |  |  |  |  |
| INGARCH–NB | Renewal–NB | 1 | 33 | 3.132 | 0.0017 | INGARCH–NB |
| INGARCH–NB | BiLSTM–NB | 1 | 33 | 0.069 | 0.9452 | Tie (ns) |
| Renewal–NB | BiLSTM–NB | 1 | 33 | -3.148 | 0.0016 | BiLSTM–NB |
| Renewal+Climate (light) | BiLSTM–NB | 1 | 33 | -1.941 | 0.0522 | BiLSTM–NB (borderline) |
| **H=2 months** |  |  |  |  |  |  |
| INGARCH–NB | BiLSTM–NB | 2 | 33 | 5.338 | < 0.001 | INGARCH–NB |
| Renewal–NB | BiLSTM–NB | 2 | 33 | -3.411 | 0.0006 | BiLSTM–NB |
| Renewal+Climate (light) | BiLSTM–NB | 2 | 33 | -3.023 | 0.0025 | BiLSTM–NB |
| **H=3 months** |  |  |  |  |  |  |
| INGARCH–NB | BiLSTM–NB | 3 | 33 | -1.729 | 0.0838 | BiLSTM–NB (ns) |
| Renewal–NB | BiLSTM–NB | 3 | 33 | -3.906 | 0.0001 | BiLSTM–NB |
| Renewal+Climate (light) | BiLSTM–NB | 3 | 33 | -4.989 | < 0.001 | BiLSTM–NB |

**Notes:** Aligned samples ensure a common (*issue\_date*, *target\_date*) per horizon ( $n = 33$ ). “ns” denotes not significant at 0.05. This subset highlights head-to-head comparisons that underpin the main claims: (i) parity between BiLSTM–NB and INGARCH–NB at $h = 1$ ; (ii) INGARCH–NB superiority at $h = 2$ ; and (iii) BiLSTM–NB superiority over mechanistic renewal baselines at $h = 3$ .

##### i) Horizon *h* = 1

INGARCH–NB significantly outperforms Renewal–NB (*t* = 3.132, *p* = 0.0017), and NB–GLM also outperforms Renewal–NB (*t* = 2.108, *p* = 0.035). INGARCH–NB and BiLSTM–NB are statistically indistinguishable at this horizon (*t* = 0.069, *p* = 0.945), consistent with their nearly equal mean log scores in the aligned summary. Relative to mechanistic baselines, BiLSTM–NB is significantly better than Renewal–NB (*t* = −3.148, *p* = 0.0016); the comparison vs. Renewal+Climate (light) is borderline at the 5% level (*t* = −1.941, *p* = 0.052).

##### ii) Horizon *h* = 2

INGARCH–NB is significantly better than BiLSTM–NB (*t* = 5.338, *p <* 0.001), matching the ranking in the aligned table. Against mechanistic baselines, BiLSTM–NB significantly improves over Renewal–NB (*t* = −3.411, *p* = 0.0006) and Renewal+Climate (light) (*t* = −3.023, *p* = 0.0025).

##### iii) Horizon *h* = 3

BiLSTM–NB significantly outperforms both Renewal–NB (*t* = − 3.906, *p* = 0.0001) and Renewal+Climate (light) (*t* = − 4.989, *p <* 0.001). The INGARCH–NB vs. BiLSTM–NB comparison is not statistically significant at the 5% level (*t* = − 1.729, *p* = 0.084), despite BiLSTM–NB attaining the best mean log score in the aligned summary at *h* = 3.

##### iv) Role of climate covariates

Within the selected head–to–heads, the Renewal+Climate (light) variant does not overturn the main conclusions: BiLSTM–NB remains significantly better than Renewal+Climate at *h* = 2–3, and parity between INGARCH–NB and BiLSTM–NB persists at *h* = 1. Overall, the DM results corroborate the aligned performance table: INGARCH–NB leads at *h* ≤ 2, while BiLSTM–NB is competitive at *h* = 1 and significantly outperforms mechanistic renewal baselines at *h* = 2–3, achieving the top mean log score at *h* = 3.

#### Forecast Calibration Diagnostics

We assess probabilistic calibration by comparing empirical predictive–interval coverage against nominal targets and by inspecting interval sharpness by horizon. Figure 3 plots empirical coverage versus nominal levels; a perfectly calibrated model lies on the 45° line. Figure 4 summarizes median interval widths (50% and 90%) by model for each horizon.

**Fig 3.**
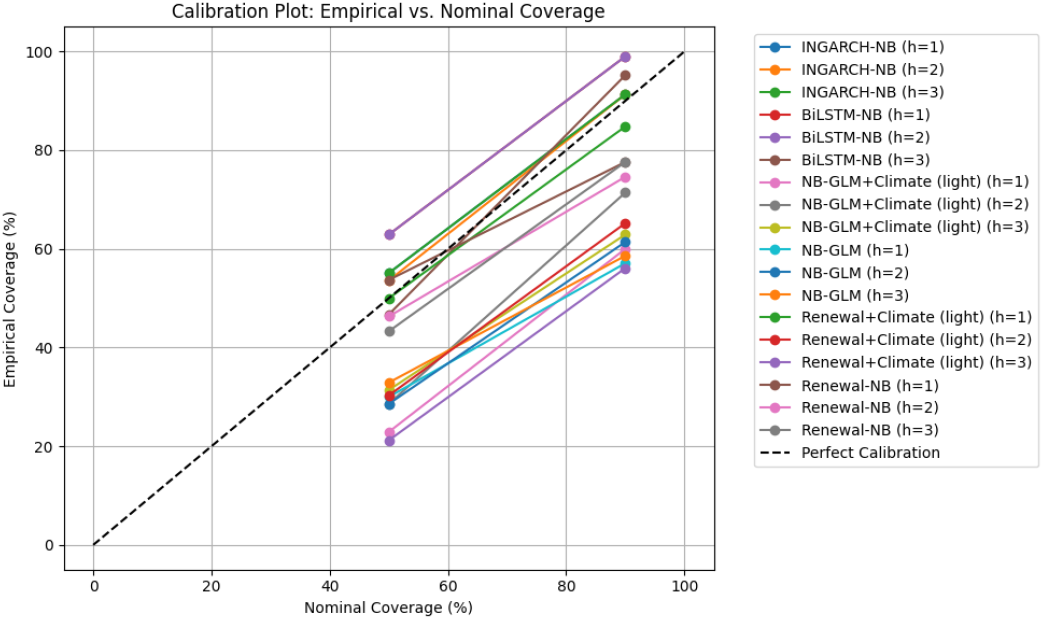
Calibration plot: empirical vs. nominal coverage. Each point shows empirical coverage at a nominal level for a given model and horizon (*h* = 1, 2, 3). The dashed line indicates perfect calibration. BiLSTM–NB generally lies above the line at 90% (slightly conservative), INGARCH–NB is close to nominal at *h* = 1 and *h* = 2, while NB–GLM variants under-cover.

**Fig 4.**
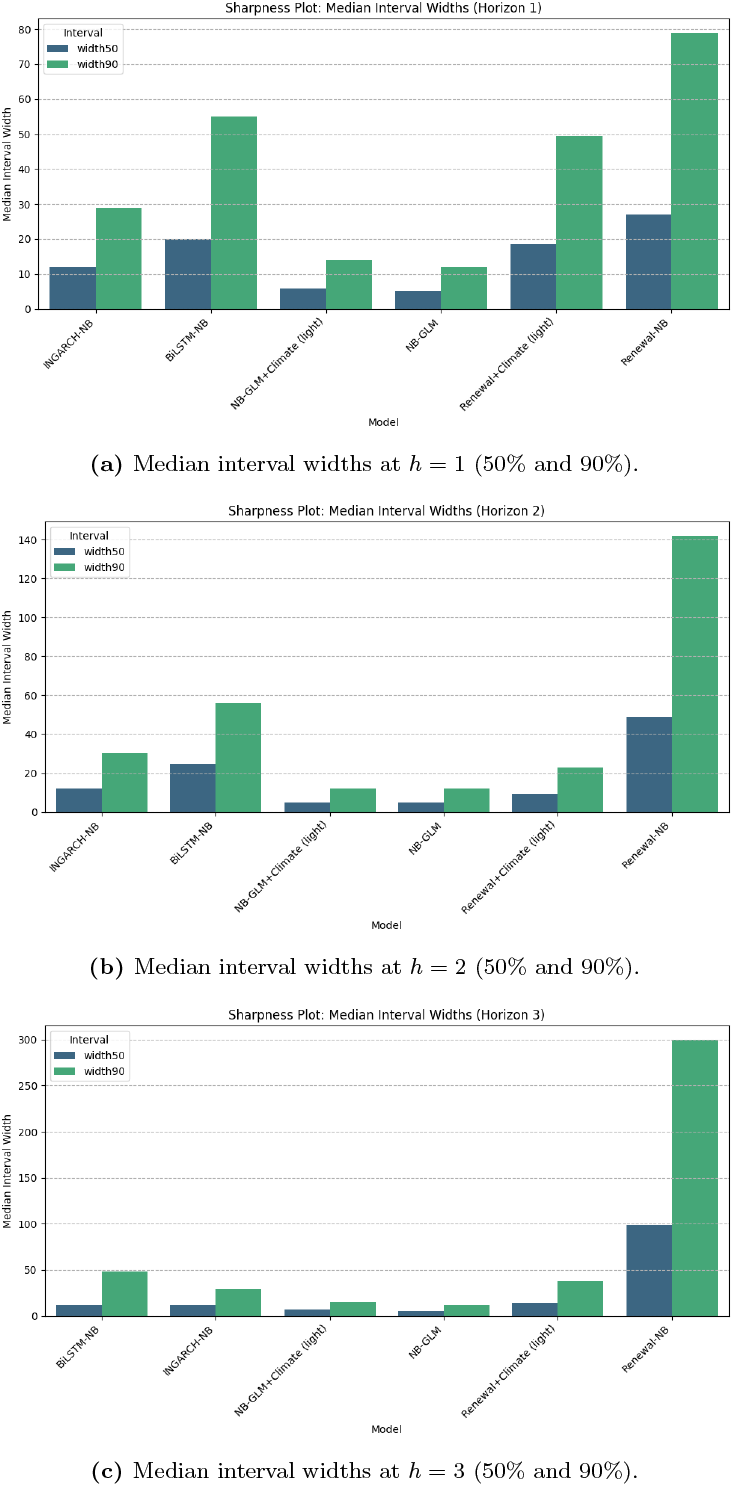
Sharpness diagnostics: median interval width by model and horizon. Narrower is better, conditional on adequate calibration. NB–GLM variants are sharpest but under-cover; BiLSTM–NB attains good coverage with moderate widths; Renewal–NB is widest, especially for *h* = 2–3.

##### Empirical vs. nominal coverage

Across aligned samples, BiLSTM–NB delivers consistently high 90% coverage (≈ 97% at all *h*), indicating slightly conservative intervals; points lie above the diagonal at 90% (Figure 3). INGARCH–NB is close to nominal at *h* = 1 (88–90%) and slightly over-covering at *h* = 2 (∼ 97%), with mild under-coverage at *h* = 3 (88–89%). In contrast, NB–GLM and its light–climate variant, under-cover at both 50% and 90% (points below the diagonal), reflect overly narrow intervals. The mechanistic Renewal–NB variants are generally over-dispersed at longer horizons (coverage above nominal), consistent with wide intervals.

##### Sharpness (interval width)

Sharpness patterns in Figure 4 mirror the coverage trade-off. NB–GLM (and NB–GLM+Climate) yields the narrowest 50%/90% intervals but under-covers, especially at *h* = 2–3. BiLSTM–NB achieves good calibration with moderate widths (e.g., median *W*_90_ ≈ 48 at *h* = 3), while INGARCH–NB is comparable in width to BiLSTM at *h* = 1 and slightly wider at longer horizons. Renewal–NB produces the widest intervals at *h* = 2–3, consistent with its conservative coverage. Overall, these diagnostics indicate that the deep model attains reliable coverage without excessively sacrificing sharpness, whereas purely GLM approaches are sharp but under-dispersed.

#### Seasonal Performance Analysis

To examine performance across the seasonal cycle, we computed the mean log score by calendar month for top models on aligned sets. Figure 5 shows BiLSTM-NB versus INGARCH-NB at horizons *h* = 1, 2, 3.

**Fig 5.**
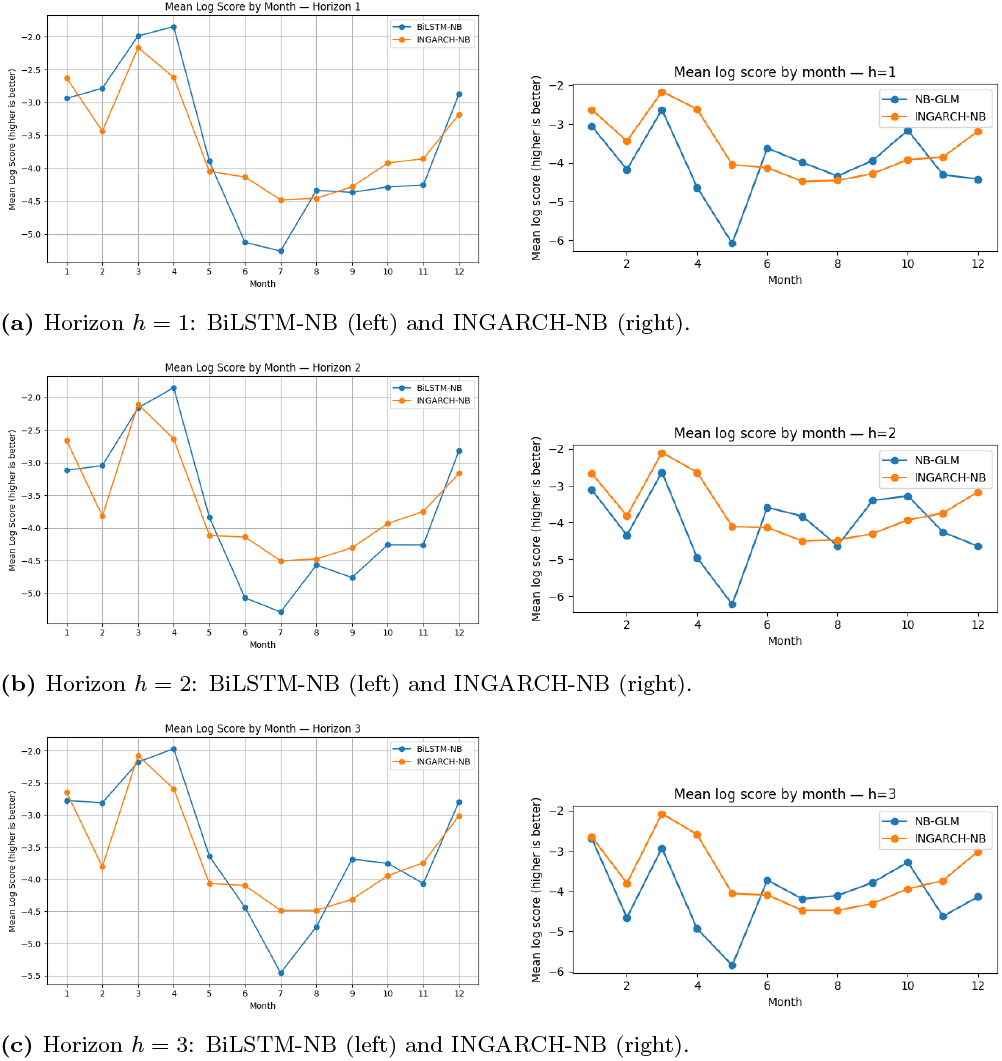
Seasonal breakdown of mean log score by calendar month. Higher (less negative) is better. Both models perform best in months with smoother dynamics; BiLSTM-NB shows increasing advantage at *h* = 3.

##### Findings

Both models tend to achieve their best (least negative) mean log scores in late dry/early rainy months (around March–May), when incidence patterns exhibit smoother dynamics. Performance degrades during mid-year peaks and transitions, where abrupt changes and outbreak spikes reduce predictability. At *h* = 1, BiLSTM-NB and INGARCH-NB are comparable month-to-month; at *h* = 2 the INGARCH-NB advantage observed in the aggregate persists in several mid-year months; at *h* = 3 the BiLSTM-NB more often leads, consistent with its overall horizon-3 superiority in Table 1. These month-resolved results reinforce that (i) short-horizon predictability is high near seasonal plateaus for both models, and (ii) representation learning confers robustness to longer-horizon drift for BiLSTM-NB.

#### Illustrative Forecasts

To provide an intuitive view of probabilistic performance, we present fan charts (90% central predictive intervals with median trajectories) for the two strongest models in our study: INGARCH–NB and BiLSTM–NB. Figure 6 displays 1-, 2-, and 3-month ahead forecasts from the INGARCH–NB model; Figure 7 shows the corresponding horizons for the BiLSTM–NB model. In all panels, the dashed line denotes observed monthly cases. Overall, the medians track intra-annual rises and declines well, particularly around peak seasons. The INGARCH–NB intervals tend to be narrower at short horizons while remaining reasonably well calibrated, whereas BiLSTM–NB intervals are typically wider and capture a larger share of peak variability, especially at *h* = 3. These patterns are consistent with the coverage and sharpness results in Figure 3-4.

**Fig 6.**
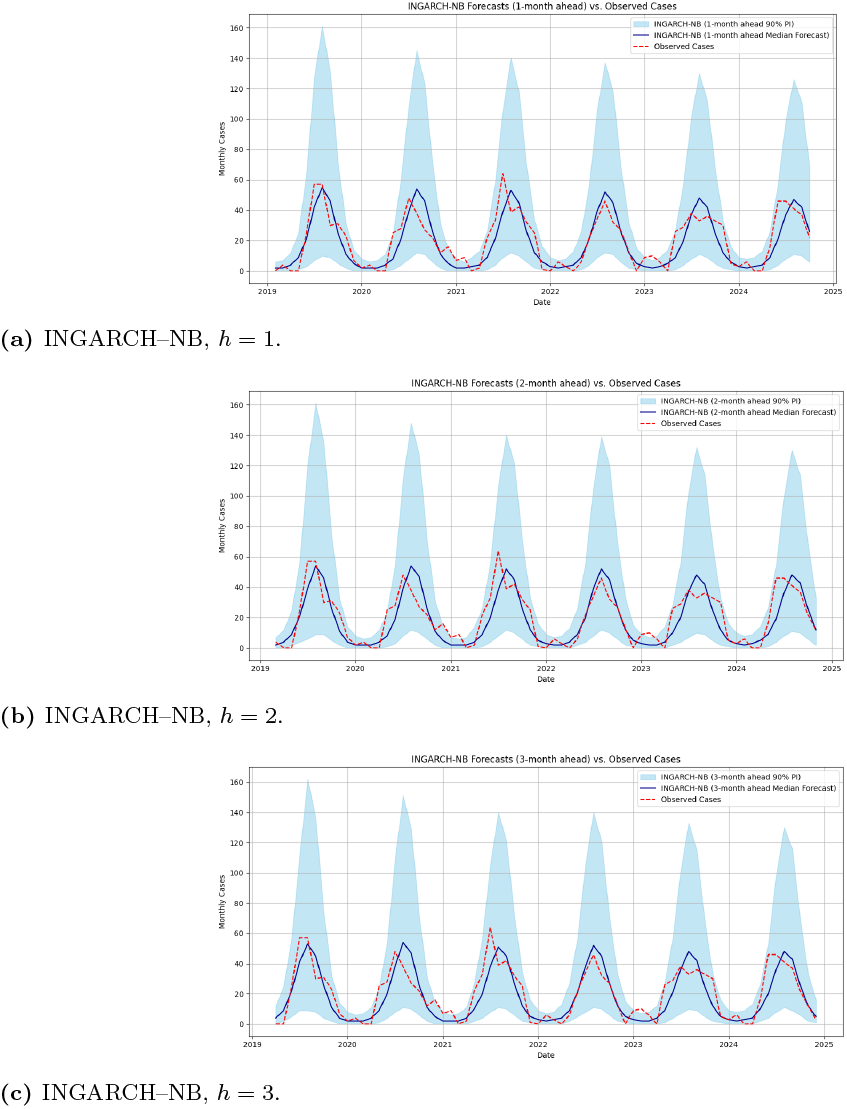
Illustrative probabilistic forecasts (fan charts) versus observed dengue cases. Shaded bands show 90% central prediction intervals; solid lines show the model median; dashed lines show observed cases. INGARCH–NB exhibits tighter fans and strong near-term tracking.

**Fig 7.**
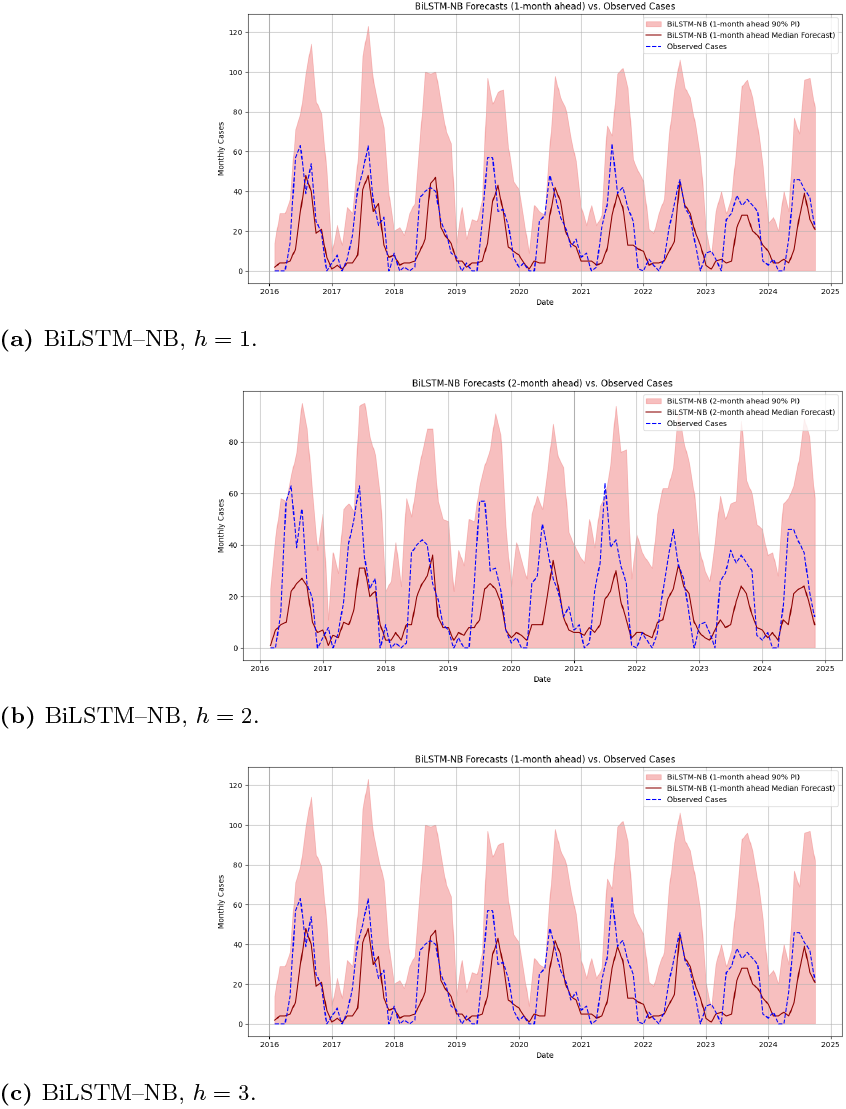
Illustrative probabilistic forecasts (fan charts) versus observed dengue cases. Shaded bands show 90% central prediction intervals; solid lines show the model median; dashed lines show observed cases. BiLSTM–NB provides broader uncertainty that better accommodates peak variability at longer horizons.

## Discussion

Resource-constrained settings increasingly use forecasts of infectious disease burden to guide preparedness, triage vector control, and communicate risk. Using monthly dengue surveillance from Freetown (2015–2025), we evaluated a spectrum of probabilistic forecasting approaches, statistical count models (NB–GLM, INGARCH–NB), a mechanistic renewal model with NB observation, and a deep learning model (BiLSTM–NB) under a harmonized expanding-window, rolling-origin design. Three findings with direct relevance to global public health emerged. First, short-lead accuracy (1–2 months) was highest for INGARCH–NB, which couples autoregressive dependence with parsimonious dynamics. Second, longer-lead accuracy (3 months) and 90% coverage reliability favored BiLSTM–NB, particularly when endemic–epidemic transitions and seasonal shoulders complicate extrapolation. Third, purely mechanistic renewal models underperformed at monthly cadence with the short serial kernel used here, emphasizing the cost of misspecifying generation time structure when data are coarse or noisy.

### Implications for operational decision–making

Public health programs typically balance *near-term responsiveness* (procurement, clinical readiness) with *medium-range planning* (larval source management, community engagement before seasonal upswings). Our results support a *portfolio* deployment: INGARCH–NB for *h* ≤ 2, where tight, sharp forecasts are valuable for immediate actions, and BiLSTM–NB for h=3, where better reliability (close-to-nominal 90% coverage) is preferable to excessive sharpness that risks underestimation. The differing strengths across horizons argue against a single “best” model and in favor of horizon-specific ensembles or simple switching rules embedded in early warning systems. In practice, programmatic triggers (e.g., “initiate pre-peak vector control if the 90% lower bound exceeds threshold X”) should be calibrated to the empirical coverage we report, not to idealized nominal levels.

### Climate, seasonality, and feasibility

We restricted exogenous inputs to “light” climate features (≤ 3) and enforced leakage safety (lag-1 only for BiLSTM; fitting only for renewal “+Climate”). This is operationally important where real-time climate feeds are uncertain or delayed. Even under these constraints, seasonality captured by harmonic terms provided a substantial signal, and climate features modestly improved some baselines without changing the qualitative ranking. For deployment in other geographies, richer meteorological and entomological drivers (humidity anomalies, vector indices) could be incorporated if and only if real-time availability and latency are validated. Otherwise, leakage and missingness risks may outweigh gains.

### Equity and systems integration

In many low- and middle-income settings, forecasting systems must interoperate with routine surveillance of variable quality. The methods used here emphasize (i) *calibration monitoring* (coverage, PIT) to avoid overconfident recommendations; (ii) *simplicity where possible* (INGARCH–NB and NB–GLM are easy to maintain, audit, and transfer); and (iii) *transparent outputs* (fan charts, aligned evaluation) that can be communicated to non-technical stakeholders. These features facilitate equitable uptake by district teams with limited analytics capacity. For sustained impact, models should be embedded within local data pipelines, accompanied by training, and augmented with human-in-the-loop review to adjudicate discordant signals.

### Interpretability versus performance

Mechanistic renewal models remain attractive because *R*_*t*_ has a clear epidemiological meaning and can inform targeted control. However, at a monthly cadence, a short, fixed serial kernel may misrepresent transmission timing; this likely contributed to underperformance and excessively wide intervals. Statistical and deep models, while less interpretable, adapted better to observed dependence structures. A pragmatic compromise is to report both: operational forecasts from the best statistical/deep model and a companion mechanistic *R*_*t*_ series for situational awareness. Where mechanistic interpretability is paramount, richer kernels (data-informed serial intervals), hierarchical pooling across years, or semi-mechanistic embeddings into RNNs may close the gap.

### Comparison with prior work

Our horizon-dependent ranking aligns with broader evidence: autoregressive count models excel at short leads; recurrent networks and hybrid architectures gain advantage as the forecast horizon grows and nonlinearity accumulates; and mechanistic models require careful specification and high-resolution data to be competitive. Importantly, our evaluation used aligned issue–target pairs and leakage controls, addressing common pitfalls that can inflate claimed gains from exogenous covariates or complex architectures.

### Strengths and limitations

Strengths include (i) a unified probabilistic scoring framework (mean log score), (ii) explicit calibration diagnostics (coverage targets and PIT), (iii) leakage-safe climate handling, and (iv) aligned backtests to ensure fair comparisons. Limitations are notable. First, analyses are confined to a single city and a monthly cadence; generalizability across space, serotype landscapes, and surveillance regimes remains to be established. Second, “light” climate inputs may underutilize environmental information; however, our finding reflects a deliberate design choice for real-time feasibility. Third, we did not model reporting delays or structural breaks (e.g., COVID-19 disruptions); in practice, adaptive schemes (rolling reweighting, change-point detection) are advisable. Fourth, while the BiLSTM–NB used isotonic calibration (first 60% of issues), alternative recalibration strategies (e.g., quantile mapping) may further improve reliability.

### Guidance for scale–up

For public health agencies considering implementation, we propose the following: (1) adopt a two-tier tool—INGARCH-NB for *h* ≤ 2, BiLSTM-NB for h=3 with automated horizon-specific selection; (2) operationalize weekly or biweekly forecast cycles synced to decision calendars; (3) monitor calibration online with rolling coverage/PIT dashboards and trigger retraining if deviations persist; (4) predefine action thresholds using retrospective coverage to mitigate overconfidence; and (5) establish data governance for climate feeds (latency audits, backfill policies) before expanding exogenous inputs. These steps align methodological rigor with institutional capacity, a prerequisite for equitable global public health deployment.

### Future directions

Methodologically, three avenues are promising: (i) *hybrid* renewal–RNN models that retain *R*_*t*_interpretability while learning residual structure; (ii) *probabilistic ensembling* across model classes to exploit complementary strengths; and *hierarchical* sharing across districts to improve data efficiency. Substantively, integrating vector surveillance, mobility, and high-resolution climate nowcasts could extend skill, provided leakage safeguards remain central. Finally, prospective evaluations (silent trials, decision impact studies) should accompany roll-out to ensure that forecast use improves outcomes without exacerbating inequities.

In summary, our study demonstrates that carefully evaluated, leakage-safe probabilistic forecasts can provide actionable guidance for dengue preparedness. A horizon-aware portfolio can enhance early warning in urban settings like Freetown by utilizing statistical counts for near-term responsiveness and calibrated deep learning for longer-range planning. Broader validation and responsible systems integration are the next steps to realize this potential in global public health practice.

## Conclusion

We developed and rigorously compared probabilistic dengue forecasting models for Freetown, Sierra Leone (monthly cadence, 2015–2025), spanning statistical count models (NB–GLM, INGARCH–NB), a mechanistic renewal model (Renewal–NB), and a deep sequence model (BiLSTM–NB) under a leakage–safe, expanding–window, rolling–origin design. Using aligned evaluation sets and proper scores, we found that short-term performance favored classical autoregressive structure (INGARCH–NB at *h* ≤ 2), while the BiLSTM–NB achieved the strongest accuracy at *h* = 3 and produced well-calibrated uncertainty after isotonic post-processing, as confirmed by coverage and PIT diagnostics; “light” climate features *leg*3 provided modest, model-dependent gains without relying on future information. These findings suggest a pragmatic portfolio for operational early warning in resource-constrained settings: INGARCH–NB for near-term nowcasting and BiLSTM–NB for tactical, medium-range planning. Limitations include the monthly resolution, a single urban setting, and omission of spatial structure and serotype/immunity dynamics; future work should test weekly data, multicity transfer, hierarchical and hybrid (mechanistic–DL) models, and real-time pipelines linked to public health triggers. Taken together, our results demonstrate that principled probabilistic learning can augment mechanistic insight to deliver actionable, uncertainty-aware dengue forecasts for global public health.

## Data Availability

All data underlying the findings are fully available without restriction. The curated monthly dengue case series for Freetown, Sierra Leone (2015–2025), together with the monthly climate aggregates (rainfall, air temperature, relative humidity) used in this study, are deposited on Kaggle and permanently archived at the following DOI: https://doi.org/10.34740/kaggle/dsv/13257213.

## Supporting information

**S1 Fig. Model performance heatmap**. Heatmap of mean log score by (model, horizon) with annotations for empirical 50%/90% coverage; included as a compact alternative to bar charts.

**S2 Fig. PIT histograms by model and horizon (aligned samples)**. Randomized Probability Integral Transform (PIT) histograms for NB-GLM, NB-GLM+Climate (light), INGARCH-NB, Renewal-NB, Renewal+Climate (light) at *h* = 1, 2, 3.

Approximate uniformity indicates good calibration; U-shapes suggest underdispersion.

**S3 File. Reproducibility package (code)**. Scripts and notebooks to reproduce Steps 1–5 (model fitting, rolling-origin evaluation, aligned indices, DM tests, figures). Includes `step4_bilstm_nb_strong.py` and `step5_synthesis.py` with a run guide. **S1 Table. Unaligned performance summary by horizon (all available issues)**. Mean log score, empirical coverage (50%, 90%), and median widths when models are evaluated on their full available indices (varying *n*); complements the aligned main-text table.

**S4 Data. De-identified monthly dengue cases and climate aggregates (2015–2025)**. https://doi.org/10.34740/kaggle/dsv/13257213

## Acknowledgments

I gratefully acknowledge the Pan African University Institute for Basic Sciences, Technology, and Innovation for their support throughout this work. I also thank my co-authors, who served as supervisors, for their sustained guidance and mentorship during this project. Finally, heartfelt thanks go to the corresponding author’s family for their encouragement and support.

